# PREDICTION OF MORTALITY USING GET WITH THE GUIDELINE HEART FAILIURE SCORE (GWTG-HF), ITS COMPARISON WITH APACHE 2 SCORE AND ITS CORRELATION WITH NTPRO BNP LEVELS: A STUDY AMONG PATIENTS OF ACUTE DECOMPENSATED HEART FAILURE FROM SOUTH EAST ASIAN REGION

**DOI:** 10.1101/2023.05.16.23290079

**Authors:** Lalit Kumar, Himanshu Mahla, Neeraj Chaturvedi, Shashi Mohan Sharma, Rajeev Bagarhatta, V.V. Agrawal, Vijay Pathak, Chandrabhan Meena, Deepak Maheshwari, Pradeep Meena, Balbir Singh Pachar, Sohan Kumar Sharma, Dinesh Gautam, Sarita Chaudhary, Dhananjay Shekhawat, Navdeep Singh Sidhu

## Abstract

**Objectives:** To study Get with the guidelines heart failure Score (GWTG-HF) among patients of acute decompensated heart failure, correlate it with outcome of survival or death till 4 week of illness, compare its performance with APACHE 2 score and to Compare its results with NT PROBNP in predicting mortality amongst patient of south east Asian ethnicity.

**Study design:** prospective observational study at medical wards and ICUs in the Department of Cardiology, SMS medical college.

**Methods:** In this study 300 patients of acute decompensated heart failure, as per Framingham criteria were analyzed and each patient underwent a complete clinical evaluation along with calculation of GWTG HF and APACHE 2 score. All the patients had NT pro BNP test within 48 hours of admission. Patient were then followed till 4 weeks of illness for outcome of survival or death, whichever occurred earlier. GWTG HF score of the patient was then compared with APACHE 2 score and it was correlated with the outcome of patients and NT pro BNP levels.

**Results:** The GWTG-HF score could predict the outcome of survival and death in ADHF patients of south east Asian ethnicity. When compared, APACHE 2 score performed better than GWTG-HF score in predicting outcome. However, GWTG-HF score was easily calculated at the bedside and was easy to use. GWTG-HF score correlated with NT-pro BNP levels with rising score having higher NT pro BNP levels.

## INTRODUCTION

Heart failure (HF) is one of the most elusive syndromes in cardiology with high morbidity and mortality.^1,2^ Due to aging population, the number of patients with HF is predicted to increase globally ultimately leading to increased burden on healthcare systems.^3^ In order to manage patients with HF properly, including frequency of outpatient examination, optimizing doses of medications, and indications for cardiac resynchronization therapy or ventricular assist device, risk classification is a high priority. Several individual parameters like age, blood pressure, heart rate, renal function, plasma B-type natriuretic peptide (BNP) level, inflammatory markers, cytokines, echocardiographic parameters, respiratory function, anemia, and presence of sleep disordered breathing have been reported for differentiating high risk cases from low risk.^4,5,6^ Since each parameter represents only a certain aspect of HF, a comprehensive risk evaluation model is the need of the hour. Many risk stratification scores using various parameters have been reported for the prediction of all-cause mortality, sudden cardiac death, and cardiovascular events in patients with HF.^7-10^ One such example, the AHEAD (atrial fibrillation, hemoglobin, elderly, abnormal renal parameters, diabetes mellitus) score was established for long-term risk prediction in acute HF.^11^ In 2010, Peterson et al established the GWTG-HF (Get With the Guidelines–Heart Failure) risk score to predict in-hospital mortality based on a cohort of 39783 patients in 198 hospitals.^12^ Melissa Layle et. al. showed that GWTG-HF risk score, along with other previously validated HF risk scores containing similar variables, had good discrimination for in hospital and 1-year mortality in a CICU cohort of 9532 patients.^13^ Satoshi Suzuki et.al. demonstrated among Japanese population(n=1452) that the plasma B-type natriuretic peptide level significantly increased with increasing GWTG - HF risk score, also event rates of all-cause death and cardiovascular events, including worsening HF and cardiac death, significantly increased with increasing GWTG - HF risk score severity in all subjects.^14^ There is sparse data available regarding the utility of this GWTG HF score in south east Asian population^15,16^. In the present study we calculated GWTG HF score among ADHF patients admitted in our tertiary care center and correlated the scores with outcome of survival or death till 4 weeks of illness.

## MATERIALS AND METHOD

The study was approved by Institutional Ethical Committee and a written informed consent was obtained from all patients prior to their inclusion. 300 patients of acute decompensated heart failure, admitted in various wards of SMS medical college between May’21 to dec’22 was enrolled within 48 hours of admission. Patients were subjected to complete history and clinical examination. Blood investigations included all routine blood biochemistry including NT pro -BNP. GWTG-HF and APACHE2 score was calculated for each patient on enrollment. The patients were followed through a period of 4weeks. Based on the data obtained, patients were classified in to survivors and non survivors as the primary outcome. GWTG-HF score was correlated with the outcome of the patients as well their NT pro BNP levels. The GWTG-HF score was compared with APACHE 2 score to predict primary outcome.

## EXCLUSION CRIETERIA

patients who were not willing to participate in study, admitted for >48 hours in the critical care, Patient with sepsis, adrenal insufficiency, burns, and uncertain diagnosis were excluded from the study.

## STATISTICAL ANALYSIS

The data was analysed using SPSS/22.0 software. The description of quantitative variables was performed using the mean, standard deviation (SD), median and quartiles. The correlation between variables was performed using the Pearson correlation coefficient, independent t test, bi serial analysis, and linear correlation graphs. A p value of <0.05 was considered significant.

## RESULTS

The study involved 300 patients of acute decompensated heart failure of which 158(52.66%) were males and 142 (47.34%) were females (TABLE 1). The mean age of the patients in the study was 57.12±16.25 years with a range of 15-88 years. 128(42.66%) patients in the study population had LVEF of 40% or more while 172(57.33%) had LVEF of less than 40%. The 4 week in-hospital mortality rate among ADHF patients was 17.33% as 52 patients died during their illness. The mean GWTG-HF score among males was higher (57.46±10.37) as compared to females (53.67±11.3). The mortality rate was 22 (15.49%) in females and 30 (18.98%) in males suggesting males had a higher death rate as compared to females. Mean NT pro BNP levels were higher in males (4113.09±4267.78 pg/ml) than females (3834.58±4571.93 pg/ml). However, for the same GWTG-HF score females had higher NT-pro-BNP levels. APACHE 2 score were calculated in the study patients and compared to the GWTG-HF score. The GWTG-HF score correlated with outcome of patients and the death rates increased as the GWTG-HF score increased (Pearson correlation coefficient R= 0.34, p<0.00001). the GWTG-HF score performed well irrespective of the LVEF of the patient (p<0.0001). As expected, APACHE 2 score with a greater number of variables could predict the outcome better than GWTG-HF score (Pearson correlation coefficient R= 0.29 p<0.00001) Based on their GWTG-HF score, patients were organised into 7 groups with scores ranging from 0-33 (n=10), 34-50(n=84), 51-57(n=56), 58-61(n=61), 62-65(n=43), 66-70(n=28), >70(n=18). As the GWTG-HF score increased number of patients needing at least one ionotropic drug increased (TABLE 2). The mortality rates also increased with progressive increase in GWTG-HF score, it was 0 (0%),7 (8.33%), 6 (10.71%), 11(18.03%), 9(20.93%), 9(32.14%), 10(55.55%) amongst the seven groups respectively (Graph 1). The mean NT pro BNP levels among the seven groups were 833.89±140.26 pg/ml, 1578.01 ±609.1 pg/ml, 2513.514±824.12 pg/ml, 3487.31 ±1331.76 pg/ml, 4614.95±1776.60 pg/ml, 6423.07±2772.96 pg/ml, 17873. 06± 7352.97 pg/ml respectively implying that NT pro BNP levels increased with increasing GWTG-HF Score. The average GWTG-HF and NT pro BNP levels among Non survivors (63.98±10.17 and 9375.38±7155.53 pg/ml) was much higher as compared to survivors (53.92±10.33 and 2774.75± 2172.22 pg/ml).

**Table 1:** Baseline characteristics of patient

| STUDY VARIABLE | MEAN/FREQUENCY/RANGE |
| --- | --- |
| Age | 57.12±16.25 years (15-88 years) |
| Sex |  |
| Male | 158(52.66) |
| Female | 142(47.34%) |
| Total | 300 |
| MEAN<br>GWTG-HF SCORE | 55.67±10.97 |
| MEAN<br>NT-PRO BNP in pg/ml | 3981.26±4408.75 |
| 4 week MORTALITY RATE (n=52) | 17.33% |
| MEAN<br>NT-PRO BNP |  |
| 1.SURVIVORS | 2774.75± 2172.22 pg/ml |
| 2.NON SURVIVORS | 9375.38±7155.53 pg/ml |
| MEAN<br>GWTG- HF SCORE |  |
| 1.SURVIVORS | 53.92±10.33 |
| 2.NON SURVIVORS | 63.98±10.17 |
| OUTCOME AT 4 WEEKS |  |
| Survivors | 248(82.67%) |
| Non survivors | 52(17.33%) |
| MORTALITY |  |
| MALE | 30(18.98%) |
| FEMALE | 22(15.49%) |
| MEAN<br>GWTG-HF SCORE |  |
| MALE | 57.46±10.37 |
| FEMALE | 53.76±11.30 |
| MEAN<br>NT PRO BNP in pg/ml |  |
| MALE | 4113.09±4267.78 |
| FEMALE | 3834.58±4571.93 |

**Table 2:**
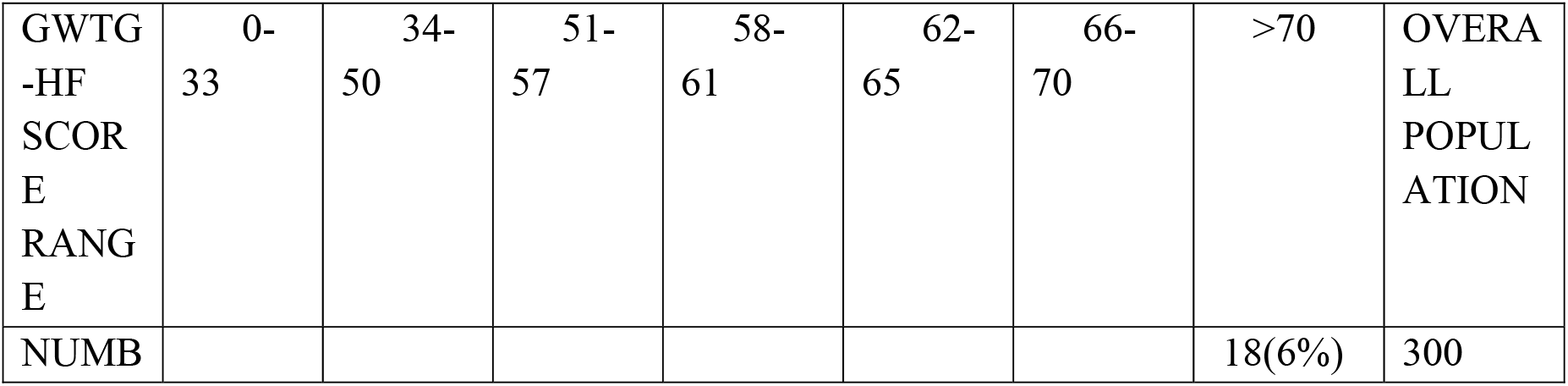
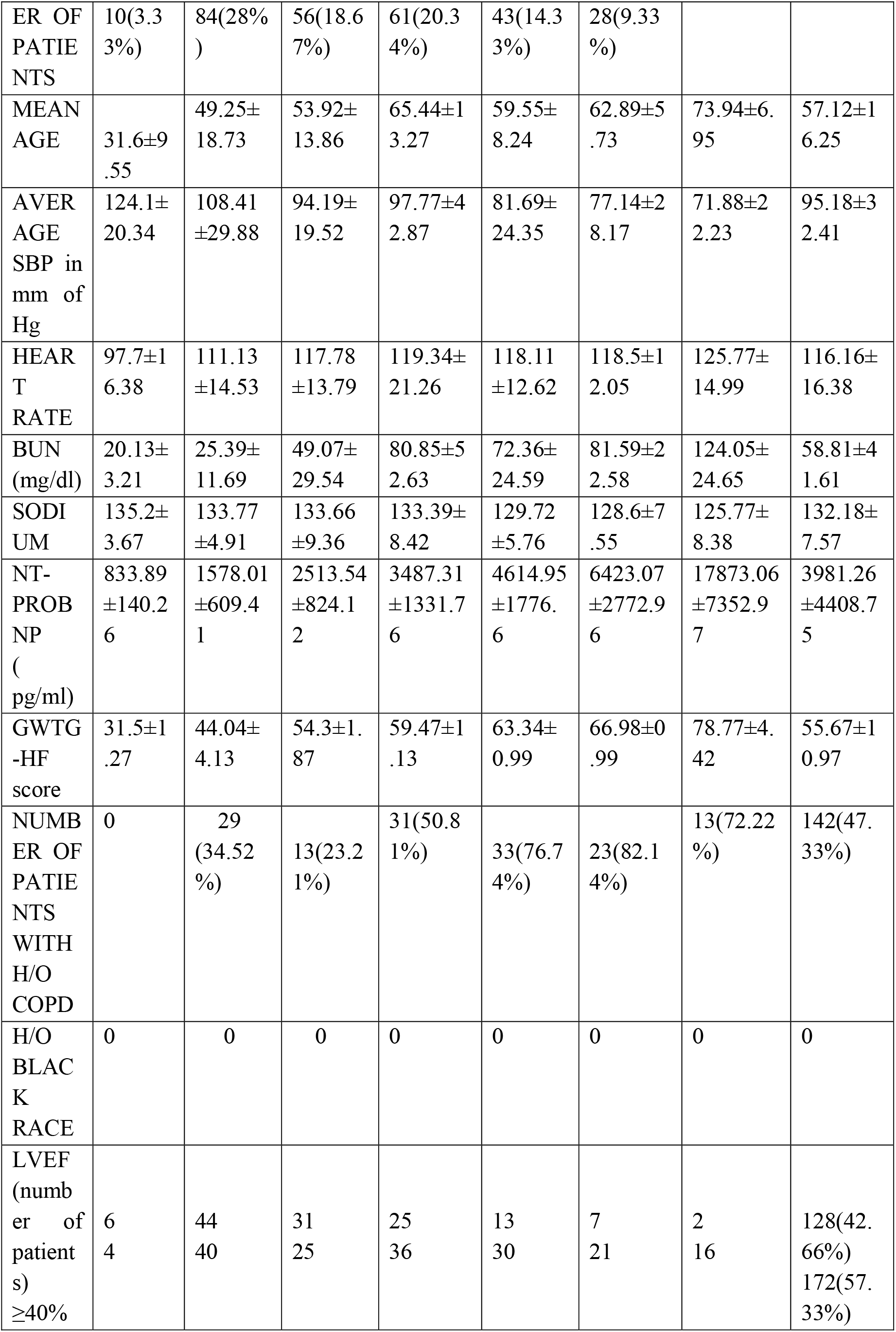
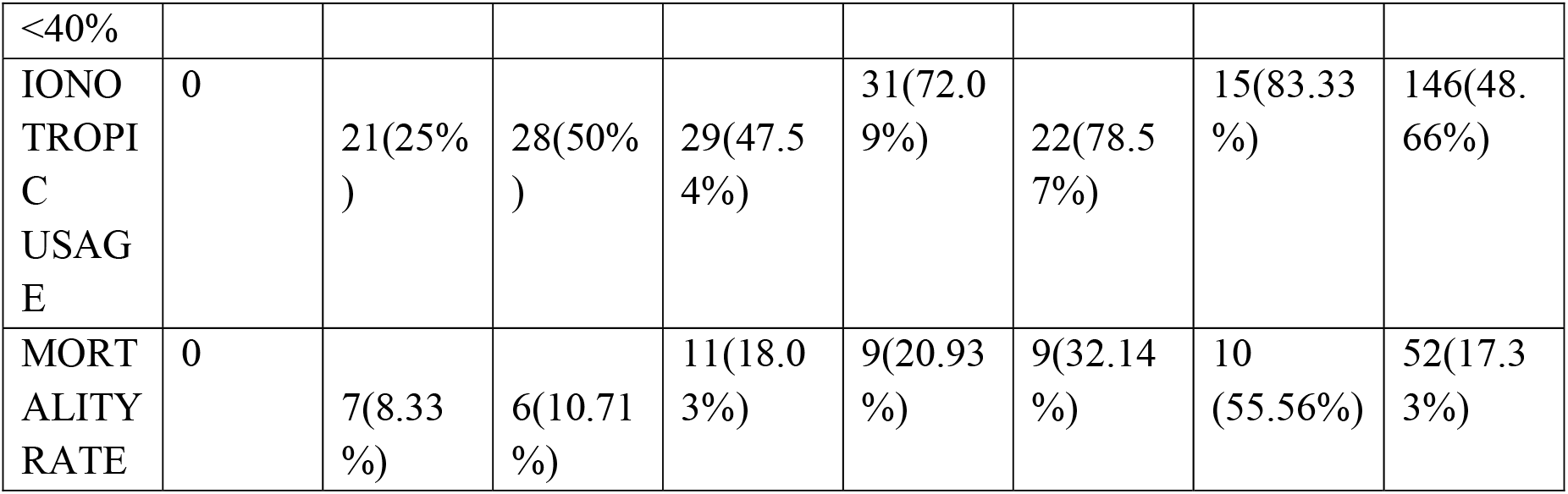
Table showing the various parameters against specified range of GWTG-HF score amongst the study population.

**Graph 1.**
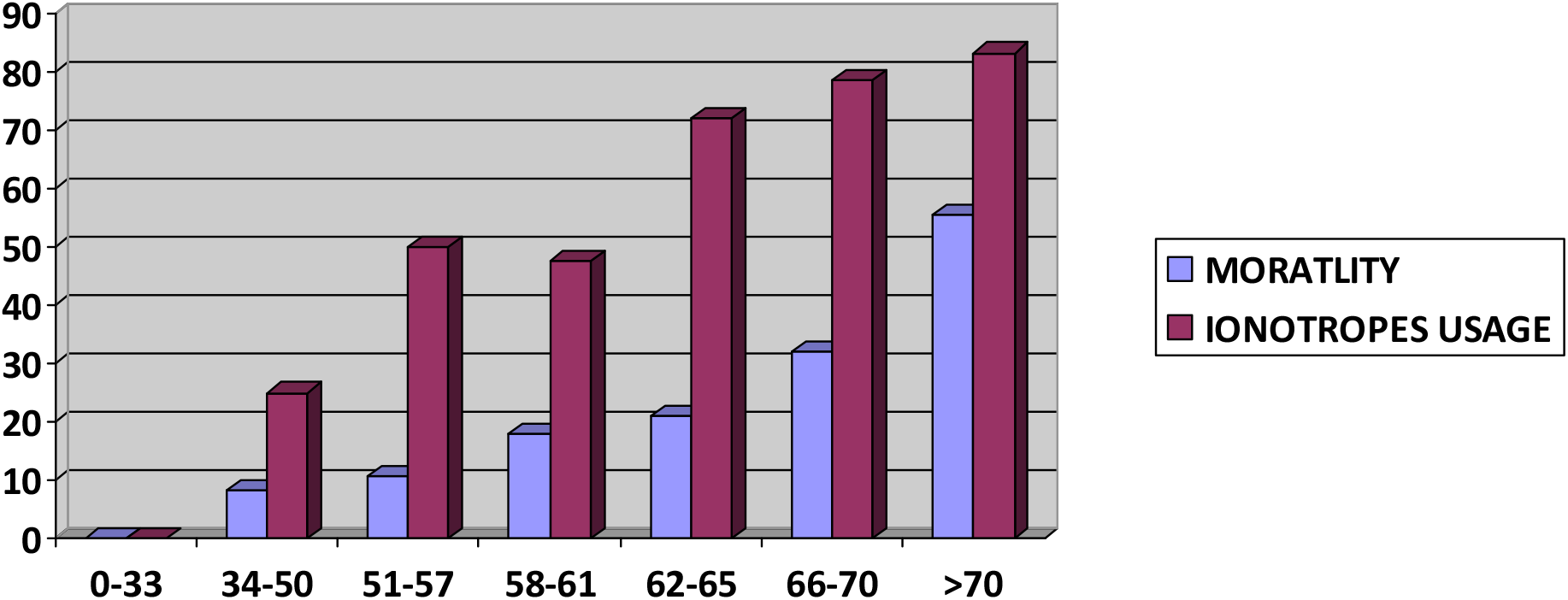
the mortality rates and use of at least 1 ionotrope in percentage (y axis) is plotted against the respective GWTG-HF score (x axis) group showing that both ionotropic use and mortality increased as GWTG-HF score increased.

The patient characteristics and study data are represented in the following tables.

## DISCUSSION

Heart failure is one of the most elusive syndromes in cardiology. Various parameters have been described for risk stratification in HF patients, but a single parameter alone is insufficient to explain the prognosis in heart failure population. Currently most patients with heart failure are elderly with many comorbidities and hence the need for a risk prediction model increase. GWTG-HF score is risk prediction score launched by AHA to prognosticate patient admitted with heart failure. GWTG-HF score is a novel risk stratification model which uses 7 variables and can be used at the bedside. The variables are Presence of Black race (points 0 or 3 point), COPD (0 or 2 point), age between 19 to 110 years (0–28 point), SBP 50 to 200 (0–28 point), BUN 9 to 150 (0–28 point), heart rate 79 to 105 (0–8 point), and serum sodium 130 to 139 (0–4 points) There are few studies amongst south east Asian population with regards to the GWTG-HF score. In this study the GWTG-HF score was studied among Indian population whose characteristics are different from the original GWTG-HF score cohort and its validity were established. The mortality rates at 4 weeks after admission in hospital were comparable and the GWTG-HF score could predict the prognosis among patients of both HFpEF and HFrEF. Heart failure mortality rates depends on intensity of observation and treatment and hence with increasing GWTG-HF score a increased level of vigilance and more intense treatment may be helpful. Amongst patient admitted with heart failure, GWTG risk score is appropriate and easy to calculate when compared with other risk prediction models. The seven variables in GWTG-HF score are routinely collected at the time of admission. After calculating GWTG-HF score, the patients can be divided in 7 groups, according to the GWTG score (0–79).

In our study, a total of 300 patient of heart failure were studied and the number of male and females in the study was comparable. Males had higher mean GWTG-HF scores and higher mortality rates, despite the fact that males had worse parameters, for the same GWTG-HF and APACHE 2 score females had higher NT-pro-BNP levels suggesting that females are somewhat protected by higher NT pro BNP levels, although other factors may also contribute. When compared, APACHE 2 score could predict the outcome better than GWTG-HF score. This is expected at it incorporates a greater number of variables which predict the outcome. However, it is cumbersome to calculate. The mean GWTG-HF score amongst the non survivors were much higher when compared with survivors. When patients were grouped according to their GWTG-HF score, most of the patients come under group 2 (28% N = 84) with GWTG score of 34 to 50 (Table 2). Mortality rates increased as the GWTG-HF score increase with scores more than 70 reaching more than 50% mortality rates. (Graph 1). the GWTG-HF score was able to predict ionotropic use among patients of heart failure with the last 3 groups having more than 50% patients who would eventually be given at-least one ionotrope during their course of hospitalization. When GWTG-HF score was correlated with NT-pro BNP levels there was a positive correlation with increasing GWTG-HF score having higher NT pro BNP values.

Among the many available heart failure risk prediction models like ADHERE, AHF INDEX, OPTIMIZE -HF SCORE every score has its own limitations. Since GWTG-HF score incorporates 7 commonly used variables which are available at the bedside it has many advantages including its wide applicability and its ability to predict mortality rates in both HFrEF and HFpEF syndromes. The GWTG-HF score predicts hospital mortality rates accurately and is a powerful tool for the treating physician. Identification of higher-risk patients sooner during their hospital course can facilitate early initiation of second-line interventions such as pulmonary artery monitoring catheters, advanced mechanical support, and even palliative care consultation. Thus, calculation of GWTG-HF score should be calculated in heart failure patients for risk quantification, triage and promotion of more invasive evidence-based therapies in subsets with high risk. There are several limitations with this study, particularly the limited sample size of only 300 and lack of racial diversity which could affect the applicability of results when applied to a broader population.

## Conclusion

In resource limited settings like those in the ASAIN countries the usage of a risk prediction score can help the physicians for accurate referral and timely intervention amongst the sickest patients of heart failure. The GWTG-HF score is based on 7 readily available parameters at the bedside and can be used to further classify the patient in a particular group and aid in clinical decision making. The better and more standardized risk assessment in the heart failure population will allow for facilitation of second-line interventions and earlier involvement of palliative care if needed. However, further studies which incorporates larger number of patients is the need of the hour.

## Data Availability

THE AUTHORS CONFIRM THAT THE DATA SUPPORTING THE FINDINGS OF THIS STUDY ARE AVAILABLE WITH IN THE ARTICLE

